# How did people cope during the COVID-19 pandemic? A Structural Topic Modelling Analysis of Free-Text Data from 11,000 UK Adults

**DOI:** 10.1101/2021.08.13.21262002

**Authors:** Liam Wright, Meg Fluharty, Andrew Steptoe, Daisy Fancourt

## Abstract

**Background:** The COVID-19 pandemic has had substantial impacts on lives across the globe. Job losses have been widespread, and individuals have experienced significant restrictions on their usual activities, including extended isolation from family and friends. While studies suggest population mental health worsened from before the pandemic, not all individuals appear to have experienced poorer mental health. This raises the question of *how* people managed to cope during the pandemic.

**Methods:** To understand the coping strategies individuals employed during the COVID-19 pandemic, we used structural topic modelling, a text mining technique, to extract themes from free-text data on coping from over 11,000 UK adults, collected between 14 October and 26 November 2020.

**Results:** We identified 16 topics. The most discussed coping strategy was ‘thinking positively’ and involved themes of gratefulness and positivity. Other strategies included engaging in activities and hobbies (such as doing DIY, exercising, walking and spending time in nature), keeping routines, and focusing on one day at a time. Some participants reported more avoidant coping strategies, such as drinking alcohol and binge eating. Coping strategies varied by respondent characteristics including age, personality traits and sociodemographic characteristics and some coping strategies, such as engaging in creative activities, were associated with more positive lockdown experiences.

**Conclusion:** A variety of coping strategies were employed by individuals during the COVID-19 pandemic. The coping strategy an individual adopted was related to their overall lockdown experiences. This may be useful for helping individuals prepare for future lockdowns or other events resulting in self-isolation.

**Correction:** Due to an error in the analytical syntax, in an earlier version of this manuscript (posted August 13, 2021), topic labels in Figure 2 were mixed up. This - and the resulting discussion - have now been corrected.

## Introduction

The COVID-19 pandemic subjected people worldwide to a range of adversities, from isolation at home to loneliness, worries about and experiences of catching the virus, troubles with finances, difficulties acquiring basic needs, and boredom (Brodeur et al., 2021; Chandola et al., 2020; ONS, 2021; Wright et al., 2020). While some of these experiences have also been reported during previous epidemics (Brooks et al., 2020), the COVID-19 pandemic was unprecedented in its global size, transmissibility, and uncertain timeframe. As a result, there were serious concerns that people would be unable to cope and there would be a substantial rise in mental illness, self-harm and suicide globally (Holmes et al., 2020; Mahase, 2020). To a certain extent, this was borne out, with data showing rises in depression and anxiety at the start of the pandemic in many countries around the world (Banks & Xu, 2020; Pierce et al., 2021). However, the COVID-19 pandemic also highlighted resilience amongst many groups, manifested as either levels of anxiety, depression and life satisfaction returning relatively quickly to pre-pandemic levels (Fancourt, Steptoe, et al., 2021; Pierce et al., 2021), or with certain groups such as older adults only experiencing small changes to their mental health (Fancourt, Steptoe, et al., 2021), or not showing any signs of worsened mental health at all (Saunders et al., 2021). This raises the question of *how* people managed to cope during the pandemic.

How people cope is an important factor underlying the relationship between experiencing stressors and subsequent mental health. Coping is generally defined as the cognitive and behavioural efforts that are used to manage stress (Lazarus & Folkman, 1991). There is much debate as to whether certain coping strategies are more beneficial than others and in what contexts. For example, strategies that aim to reduce and resolve stressors may be more effective in supporting mental health (Taylor & Stanton, 2007), but avoidant strategies may be helpful in reducing short-term stress (Taylor & Stanton, 2007). While avoidant strategies may have some benefits, they can also lead to more harm as no direct actions are taken to reduce the stressor, potentially resulting in feelings of helplessness or self-blame (Suls & Fletcher, 1985). Numerous sociodemographic, personality, and social factors are known to influence how people cope with stress (Bolger & Zuckerman, 1995; Christensen et al., 2006). For example, personality type can influence the severity of the stressor experience by facilitating or constraining use of coping strategies (Bolger & Zuckerman, 1995; Connor-Smith & Flachsbart, 2007). Additionally, effects of personality on coping are facilitated by their level of engagement with stressors, and similarly their approach to rewards (Connor-Smith & Flachsbart, 2007; Leszko et al., 2020).

A number of studies have examined coping during the COVID-19 pandemic, using quantitative (Agha, 2021; Fluharty & Fancourt, 2021; Park et al., 2020), qualitative (Ogueji et al., 2021; Sarah et al., 2021), and mixed methods approaches (Chew et al., 2020; Dewa et al., 2021). Quantitative studies have examined the association between coping strategies and the level and trajectories of symptoms of poor mental health during the pandemic (Agha, 2021; Fluharty et al., 2021). Further, a study examining predictors of coping found COVID-19 specific experiences contribute to choice of coping strategy (Fluharty & Fancourt, 2021). For instance, experience financial adversity (such as job loss and major cut in income) was association with problem-focused, emotion-focused, and avoidant coping. Additionally, two qualitative studies collected free text responses on how people were coping during the COVID-19 pandemic (Ogueji et al., 2021; Sarah et al., 2021). Both studies found the most common strategies employed were centred around socially-supported coping. However, these studies had small sample sizes and were both recruited over social media, which may have biased the sample towards this result. Further, the studies were narrative in nature and did not compare coping strategies across sociodemographic groups or in a formal manner to peoples’ lockdown experiences.

There has also been a methodological challenge with the studies on coping during COVID-19 carried out so far. Existing quantitative studies have typically relied on closed-form responses to survey items (e.g., Likert responses). An issue with this approach is that responses are restricted to those that the researcher has thought of in advance – an issue that is particularly salient given the novelty of the COVID-19 pandemic. While qualitative approaches allow for more flexibility in responses, the small sample sizes typical of qualitative studies restrict the questions that can be asked of the data – specifically, those that statistically relate individual characteristics and circumstances to topics raised.

Therefore, this study sought to fill the empirical and methodological gaps in understanding coping during COVID-19 by using structural topic modelling (STM; Roberts et al., 2014) with free-text data on coping strategies during the COVID-19 pandemic collected from 11,000 UK adults. Structural topic modelling is a text mining technique that enables the extraction of themes from large-scale open-ended free-text responses and the quantitative summary and analysis of these themes alongside other participant data (i.e., in regression models). We used STM to explore the coping strategies adopted during the pandemic, and how these strategies related to participants’ demographic, socioeconomic and personality characteristics and to their lockdown experiences.

## Methods

### Participants

We used data from the COVID-19 Social Study; a large panel study of the psychological and social experiences of over 70,000 adults (aged 18+) in the UK during the COVID-19 pandemic. The study commenced on 21 March 2020 and involved online weekly data collection for 22 weeks with monthly data collection thereafter. The study is not random and therefore is not representative of the UK population, but it does contain a heterogeneous sample. Full details on sampling, recruitment, data collection, data cleaning and sample demographics are available at https://github.com/UCL-BSH/CSSUserGuide. The study was approved by the UCL Research Ethics Committee [12467/005] and all participants gave informed consent.

A one-off free-text module was included in the survey between 14 October and 26 November 2020. Participants were asked to write responses to eight questions on their experiences during the pandemic and their expectations for the future. Here, we used responses to a single question: *What have been your methods for coping during the pandemic so far and which have been the most or least helpful?* (See Table S1 for the full list of questions). 30,950 individuals participated in the data collection containing this survey module (43.4% of participants with data collection by 26 November 2020). Responses to the free-text questions were optional. 12,536 participants recorded a response to the question on coping (40.5% of eligible participants). Of these, 11,073 (88.3%) provided a valid record, the definition of which is provided in a following section.

The period 14 October – 26 November was six months into the pandemic in the UK and overlapped with the beginning of the second wave of the virus. As such, participants could reflect on their experiences during a strict lockdown from March 2020, the relaxation of that lockdown over the summer of 2020, and the start of new restrictions being brought in for the second wave. Figure S1 shows 7-day COVID-19 caseloads and confirmed deaths, along with the Oxford Policy Tracker, a numerical summary of policy stringency (Hale et al., 2020), across the study period. An overview of the key developments in the pandemic across the study period is provided in the Supplementary Information.

### Data cleaning

We performed topic modelling using unigrams (single words). Responses were cleaned using an iterative process, described in detail in the Supplementary Information. We excluded responses containing fewer than five words and removed words which appeared in fewer than five responses. We also removed common “stop” words (“the”, “and”, “I”, etc.) from the analysis. We used complete case data so excluded a small number of participants with missing data on any covariate used in the analysis (n = 113). Spelling mistakes were identified with the hunspell spellchecker (Ooms, 2018), amended manually if they had FOUR or more occurrences, and replaced using the hunspell suggested word function if the number of occurrences was fewer than four. Where the algorithm provided multiple suggestions, the word with the highest frequency across responses was used. To reduce data sparsity, in the structural topic models, we used word stemming using the Porter (1980) algorithm. Data cleaning was carried out in R version 3.6.3 (R Core Team, 2020).

### Data analysis

We performed several quantitative analyses. First, as not all participants chose to provide a response, we ran a logistic regression model to explore the predictors of providing a free-text response. Second, we used STM, implemented with the stm R package (Roberts et al., 2019), to extract topics from responses, with the analysis carried for each question separately. STM treats documents as a probabilistic mixture of topics and topics as a probabilistic mixture of words. It is a “bag of words” approach that uses correlations between word frequencies within documents to define topics. STM allows for inclusion of covariates in the estimation model, such that the estimated proportion of a text devoted to a topic can differ according to document metadata (e.g., characteristics of its author). We included participant’s gender, ethnicity (white, non-white), age (smooth splines, degrees of freedom 4), education level (degree or above, A-Level or equivalent, GCSE or below), living arrangement (alone; alone, without child; alone, with child), psychiatric diagnosis (any, none), long-term physical health conditions (0, 1, 2+), self-isolation status (yes, no), and Big-5 personality traits (Openness to Experience, Conscientiousness, Extraversion, Agreeableness, Neuroticism), each collected at first data collection. There was only a small amount of item missingness, so we used complete case data. More detail on the variables used in this analysis is provided in the Supplementary Information.

We ran STM models from 2-30 topics and selected the final models based on visual inspection of the semantic coherence and exclusivity of the topics and close reading of exemplar documents representative of each topic. Semantic coherence measures the degree to which high probability words within a topic co-occur, while exclusivity measures the extent to that a topic’s high probability words have low probability for other topics. After selecting a final model, we carried out three further analyses. First, we decided upon narrative descriptions for the topics based on high probability words, high “FREX” words (a weighted measure of word frequency and exclusivity), and exemplar texts (responses with a higher proportion of text estimated for a given topic). Second, we ran regression models estimating whether topic proportions were related to author characteristics defined above. Third, we ran regression models estimating whether lockdown experiences were related to topic proportions, to explore which coping strategies may have been particularly effective. Lockdown experiences were measures with three separate Likert items on enjoying lockdown (How much have you enjoyed lockdown? 1. Not at all, 7. Very much), missing lockdown (Do you feel you will miss being in lockdown? 1. Not at all, 7. Very much), and feelings about future lockdowns (How do you feel about the prospect of any future lockdowns? 1. I would dread it, 7. I would really look forward to it). These variables were collected between 11-18 June 2020.

Individuals with item-missingness were dropped in each analysis. Data cleaning and analysis was carried out by LW. LW selected the number of model topics and LW and MF agreed upon narrative titles for the topics. The free-text data used in this study cannot be made publicly available due to stipulations set out by the ethics committee. The code used in the analysis is available at https://osf.io/xqu8h/.

## Results

### Descriptive Statistics

11,073 individuals provided a valid free-text response. Descriptive statistics for respondents are displayed in Table 1, with figures for the total eligible sample also shown for comparison. Participants in the COVID-19 Social Study are disproportionately female, older age, and more highly educated, relative to the general population (Fancourt, Paul, et al., 2021). There were some differences between those who provided a (valid) response and those that did not. Figure S2 displays the results of logistic regression models exploring the predictors of providing a response. Responders were disproportionately female, of older age, more highly educated, more likely to live alone, and to have self-isolated than non-responders. They were also more open, conscientious, and extraverted, on average.

**Table 1:**
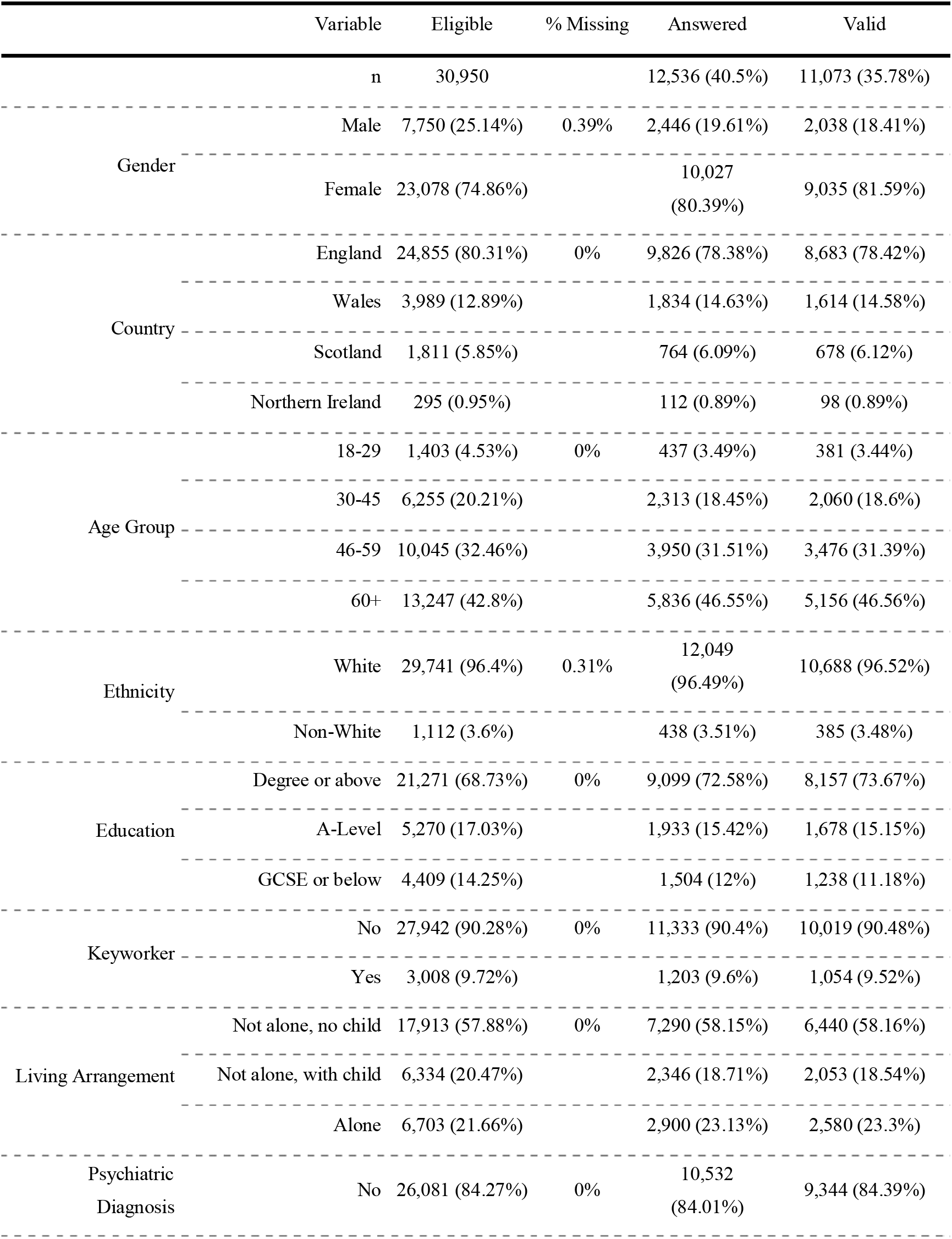
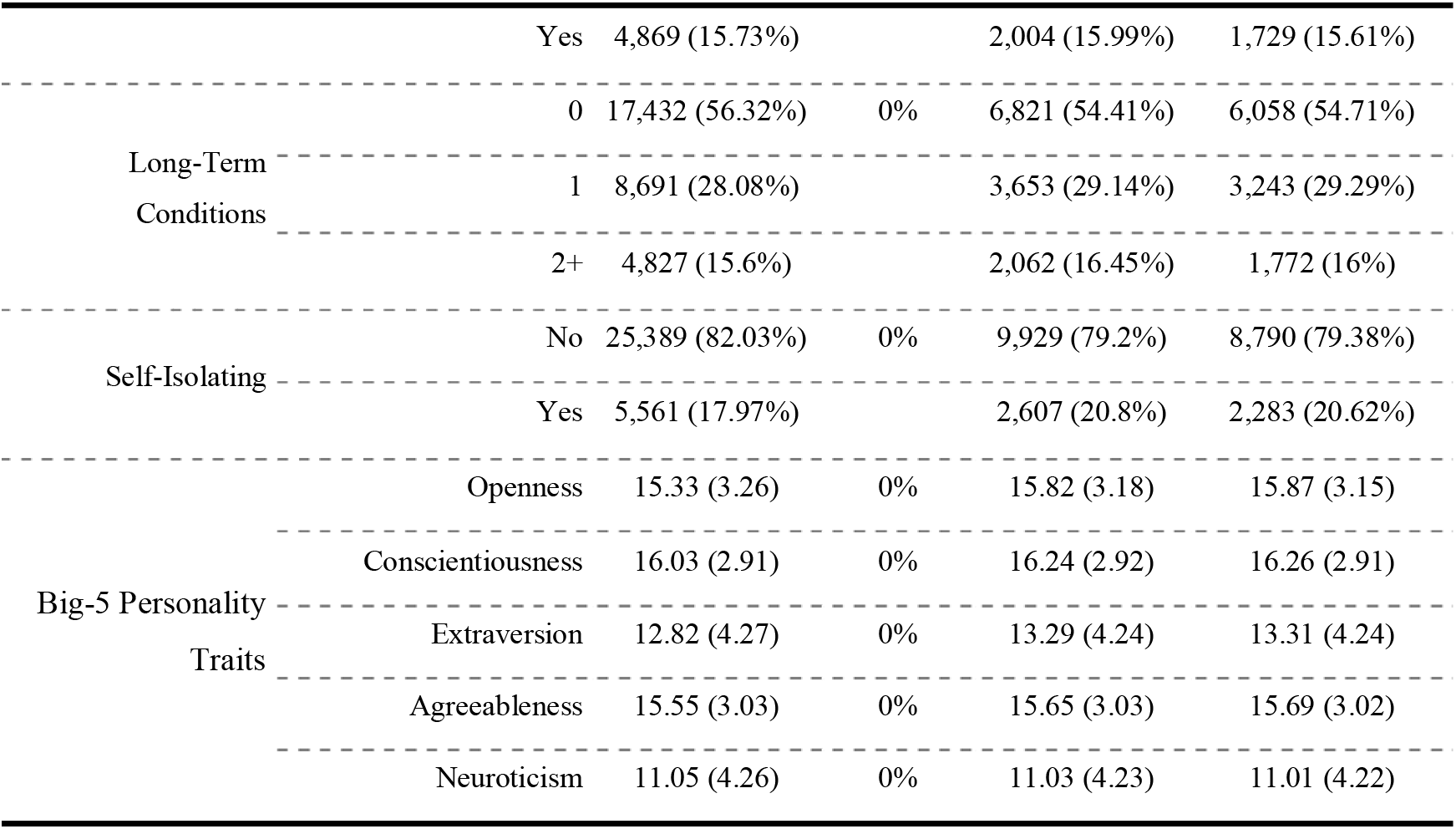
Descriptive Statistics

A word cloud of the forty most frequently used words for each question is displayed in Figure 1. Many of the words refer to activities or time use (e.g., walking, reading, exercise, routine) or to social factors (e.g., friends, family, talking, zoom).

**Figure 1:**
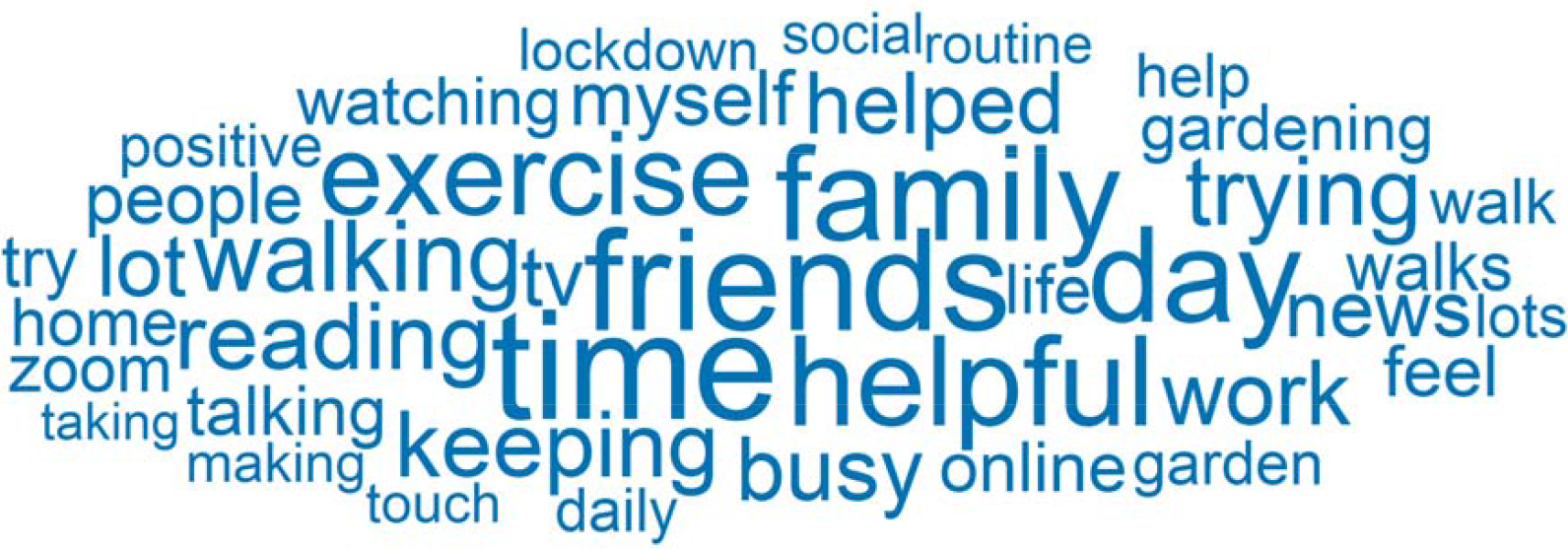
Word cloud. Forty most frequently used words across responses. Words sized according to number of responses they appear in.

**Figure 2:**
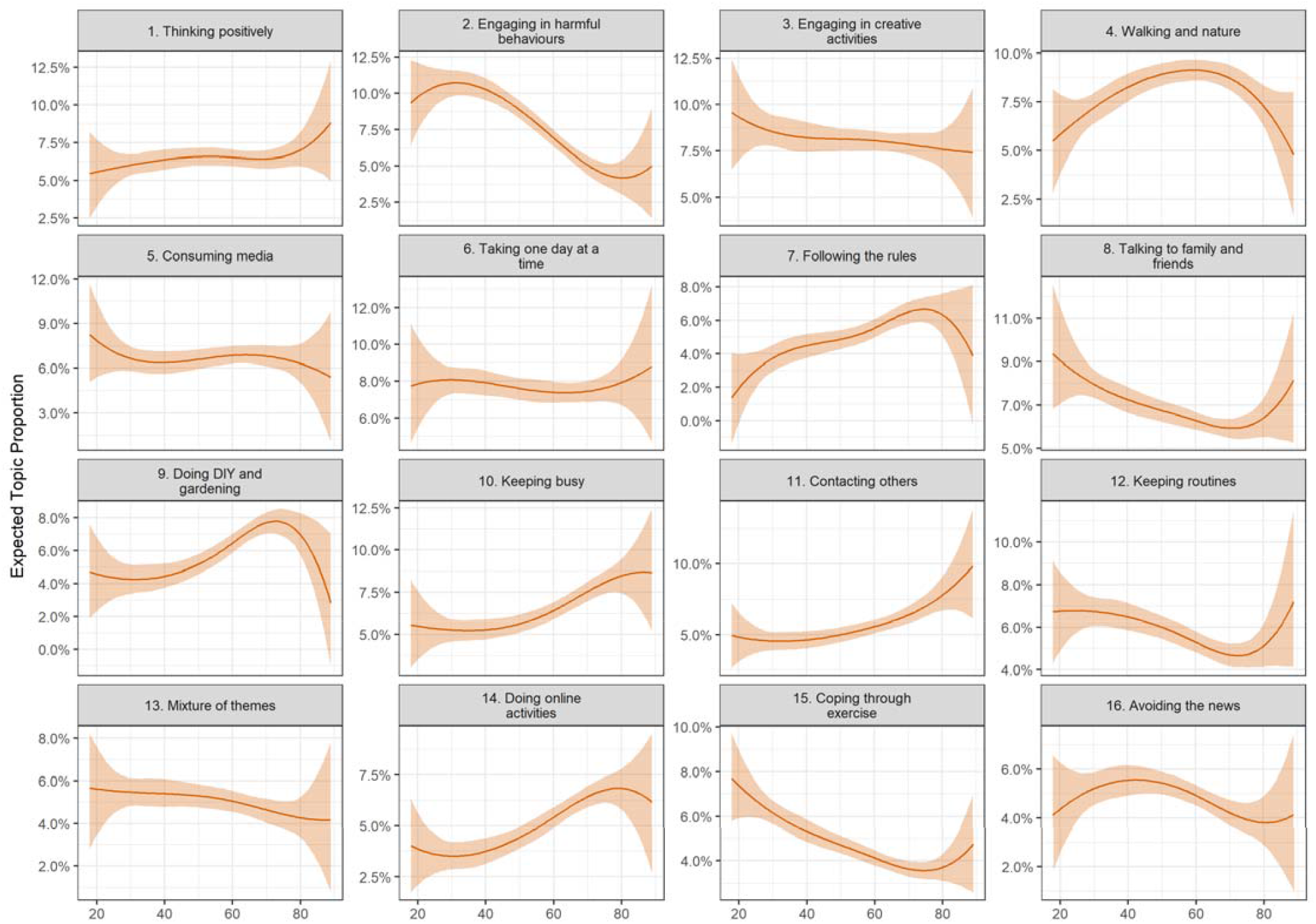
Association between document topic proportion and participant’s age (+ 95% confidence intervals). Derived from OLS regression models including adjustment for gender, ethnicity, age, education level, living arrangement, psychiatric diagnosis, long-term physical health conditions, self-isolation status, Big-5 personality traits and keyworker status.

### Coping Strategies

We selected a 16 topic solution. Short descriptions are displayed in Table 2, along with exemplar quotes and topic titles that we use when plotting results. Topics are ordered according to the estimated proportion of text devoted to each topic. Correlations between the topic proportions are displayed in Figure S3.

**Table 2:**
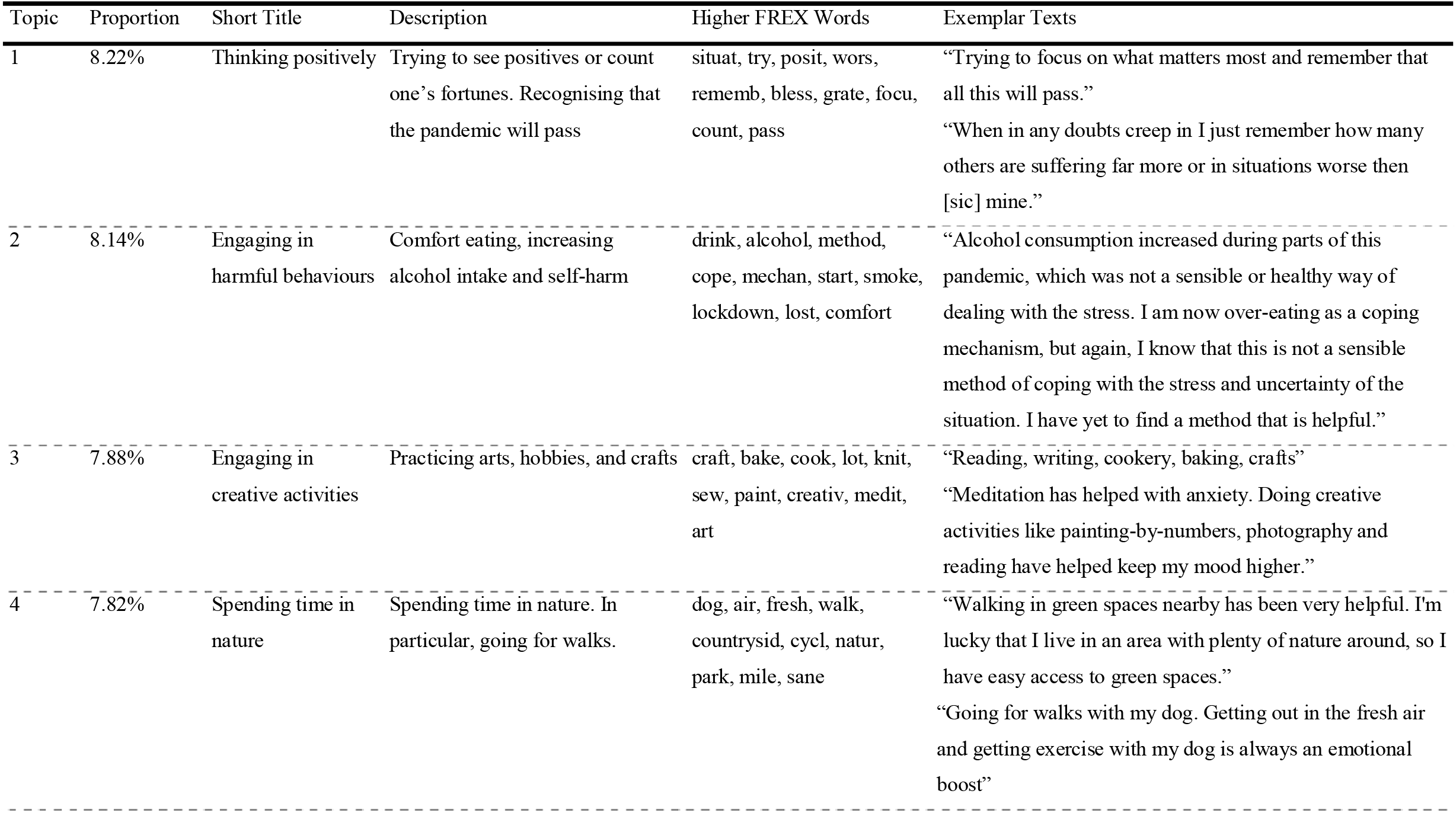
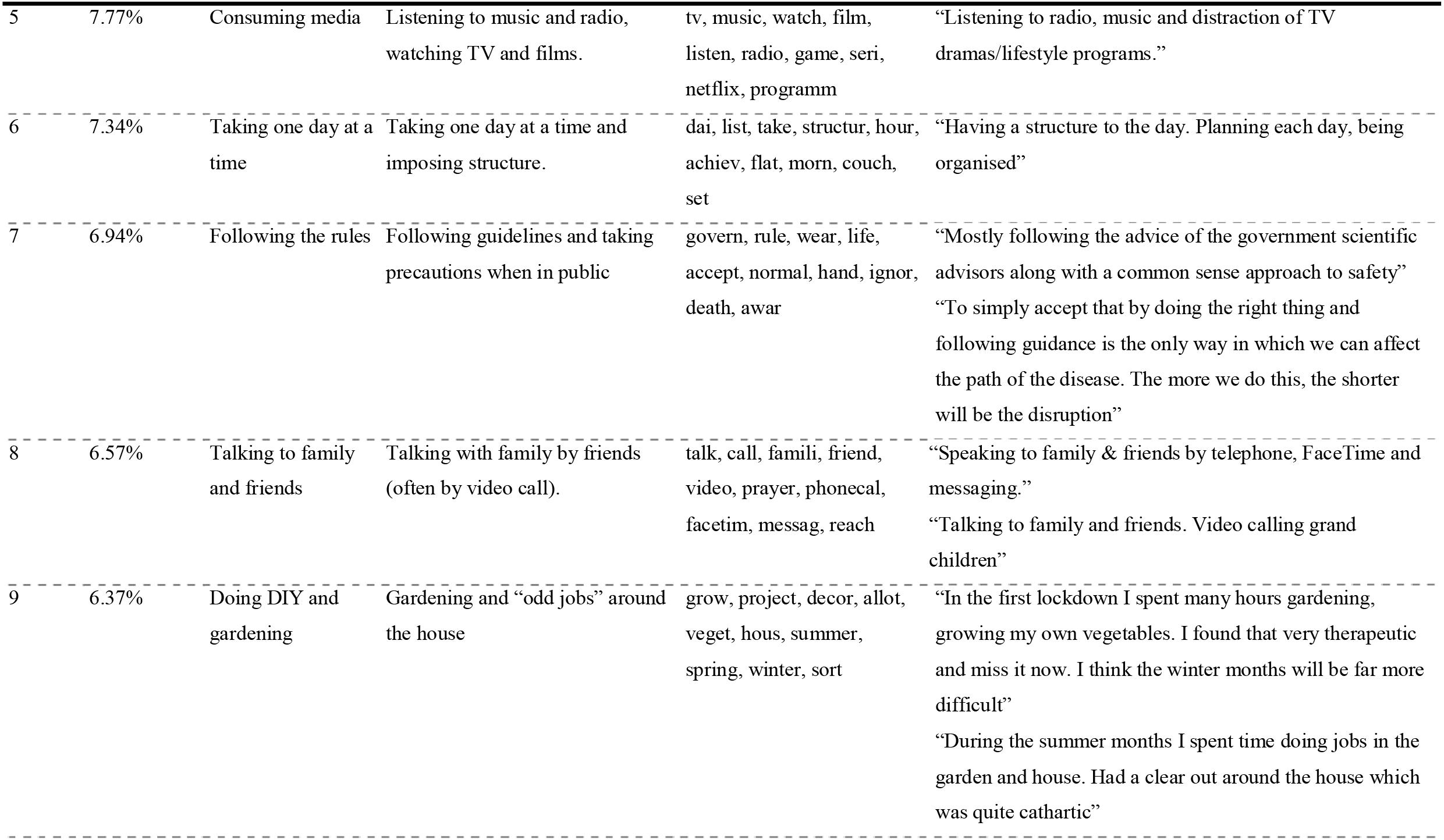
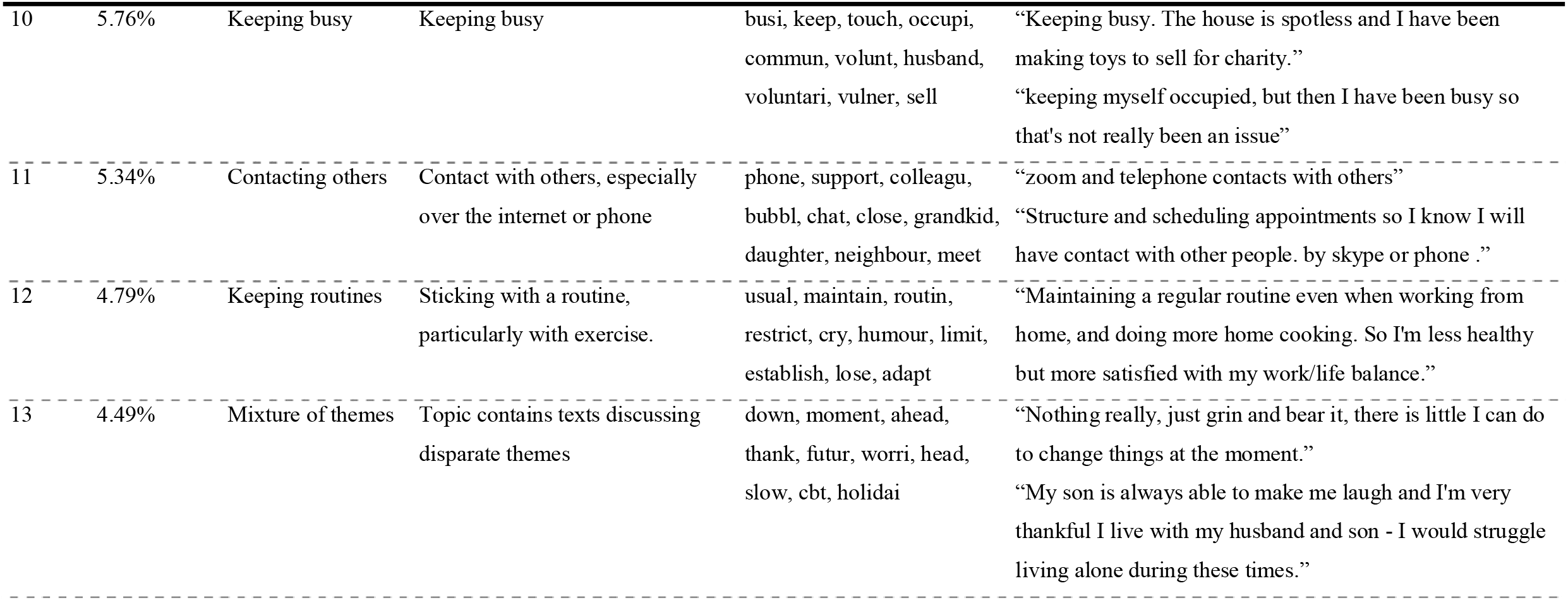
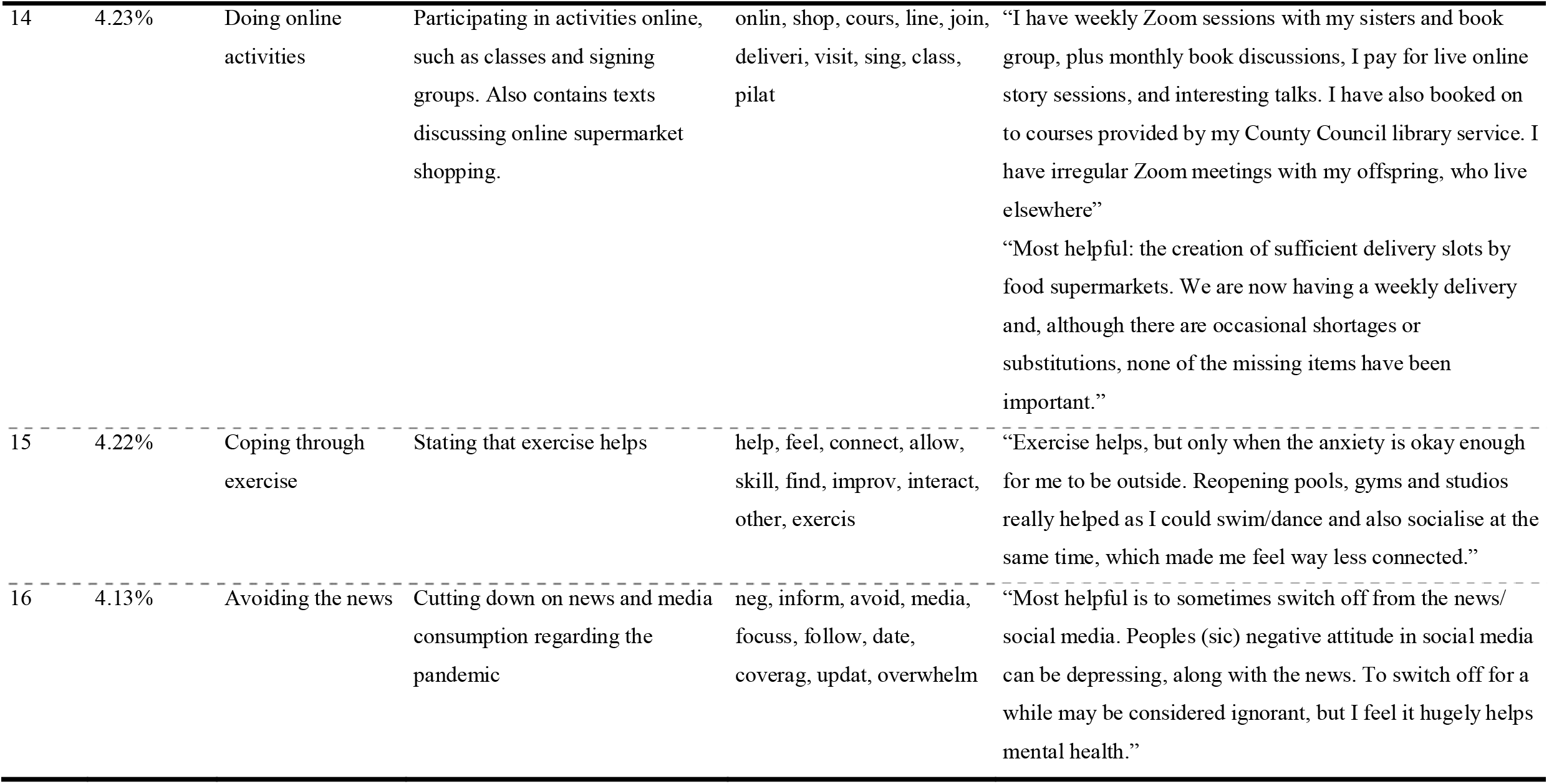
Topic Descriptions

The largest topic (Topic 1; 8.82% of text; Thinking positively) included individuals who had tried to see the positives in the situation, to remember that others were in relatively worse situations, and recognise that the pandemic would pass. Topic 6 (7.34%; Taking one day at a time) similarly related to a general cognitive coping strategy, including text on individuals taking each day as it came and imposing structure on their time. This topic overlapped with Topic 12 (4.79%; Keeping routines), which related to people keeping routines, particularly with exercise. Similarly, Topic 10 (5.76%; Keeping busy) related to participants filling their time (“keeping busy”) in generally non-specific ways.

Most other topics related to spending time on specific activities. Topics 3 (7.88%; Engaging in creative activities) related to individuals engaging in arts, hobbies, or crafts as a way of coping. Topic 5 (7.77%; Consuming media) included text from participants who reported spending their time listening to music and radio or watching TV and films. Topic 4 (7.82%; Walking and spending time in nature) related to individuals who had used the opportunity to take long walks and get into nature, while Topic 15 (4.22%; Coping through exercise) included text from individuals who found exercise had a positive effect. Topic 8 (6.57%; Talking to family and friends) and Topic 11 (5.34%; Contacting others) including responses on keeping in contact with family, friends and colleagues, the latter referring to the use of online technologies in particular. Topic 9 (6.37%; Doing DIY and gardening) referred to individuals spending time gardening or completing “odd jobs” at home. Topic 14 (4.23%; Doing online activities) included participants spending time on activities online, including classes, courses, and group sessions (such as singing groups) as well as functional online activities such as ordering supermarket deliveries.

Amongst the remaining topics, topic 2 (8.14%; Engaging in harmful behaviours) included individuals who reported self-harming or increasing alcohol consumptions or comfort eating, though the latter two were reported in several cases as improving mood (at least in the short term). Topic 16 (4.13%; Avoiding the news) meanwhile included text on individuals actively avoiding coverage on COVID-19 as a coping strategy. Topic 7 (6.94%; Following the rules) referred specifically to attempts to reduce risk by following guidelines (e.g., mask wearing). Finally, topic 13 (4.49%; Mixture of themes) surfaced exemplar texts that did not contain a clear, consistent theme.

### Topic Proportions and Author Characteristics

#### Age

When exploring associations between topic proportions and socio-demographic characteristics, there were several differences in topic proportions by age (Figure 2). Compared with middle-aged adults, younger adults were more likely to report engaging in harmful or risk behaviours (Topic 2) and coping through exercise (Topic 15). Older people, on the other hand, were more likely to discuss following the rules (Topic 7), keeping busy (Topic 10) and doing online activities (Topic 14). There were some clear non-linear associations, too – middle-aged adults were more likely to discuss walking and nature (Topic 4).

#### Personality Traits

There were also several differences according to personality traits (Figure 3). Individuals high in trait openness were more likely to engage in creative activities (Topic 3); conscientious individuals were more likely to report keeping busy (Topic 10), walking and spending time in nature (Topic 4), or spending time on DIY or gardening (Topic 9); extravert individuals were more likely to report contacting others (Topic 8 and Topic 11) and less likely to spend time consuming media (Topic 5) or doing DIY or gardening (Topic 9); agreeable individuals were more likely to avoid the news (Topic 16), spend time talking to family and friends (Topic 8) and less likely to report engaging in harmful behaviours (Topic 2); and neurotic individuals were more likely to spend time consuming media (Topic 5) and – surprisingly – less likely to report keeping routines (Topic 12) and following the guidelines (Topic 7).

**Figure 3:**
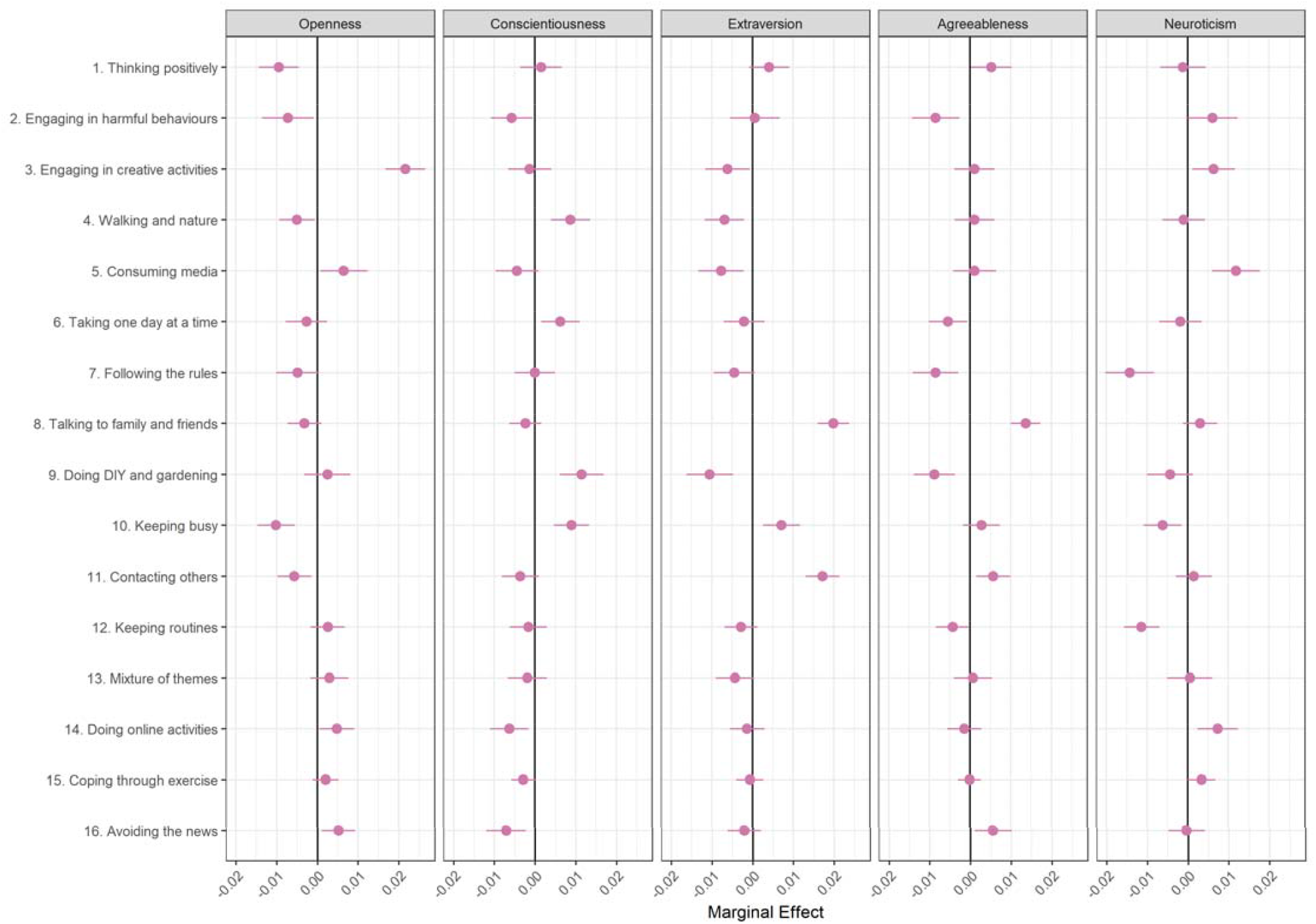
Association between document topic proportion and Big-5 personality traits (+ 95% confidence intervals). Derived from OLS regression models including adjustment for gender, ethnicity, age, education level, living arrangement, psychiatric diagnosis, long-term physical health conditions, self-isolation status, Big-5 personality traits and keyworker status.

#### Demographic, Socio-Economic, and Health Characteristics

Finally, there were also differences according to demographic, socio-economic and health characteristics (Figure 4). In particular, women were more likely to report engaging in creative activities (Topic 3) and talking with family and friends (Topic 8) and were less likely to discuss following the rules (Topic 7). Individuals with lower education were more likely to mention thinking positively (Topic 1) and following the guidelines and less likely to discuss engaging in online activities (Topic 14). Individuals who lived alone were more likely to discuss contacting others (Topic 11) and individuals with psychiatric diagnoses were more likely to report engaging in harmful behaviours (Topic 2) and less likely to report walking and spending time in nature (Topic 4). Participants in self-isolation were more likely to mention DIY and gardening (Topic 9) and engaging in creative activities (Topic 3).

**Figure 4:**
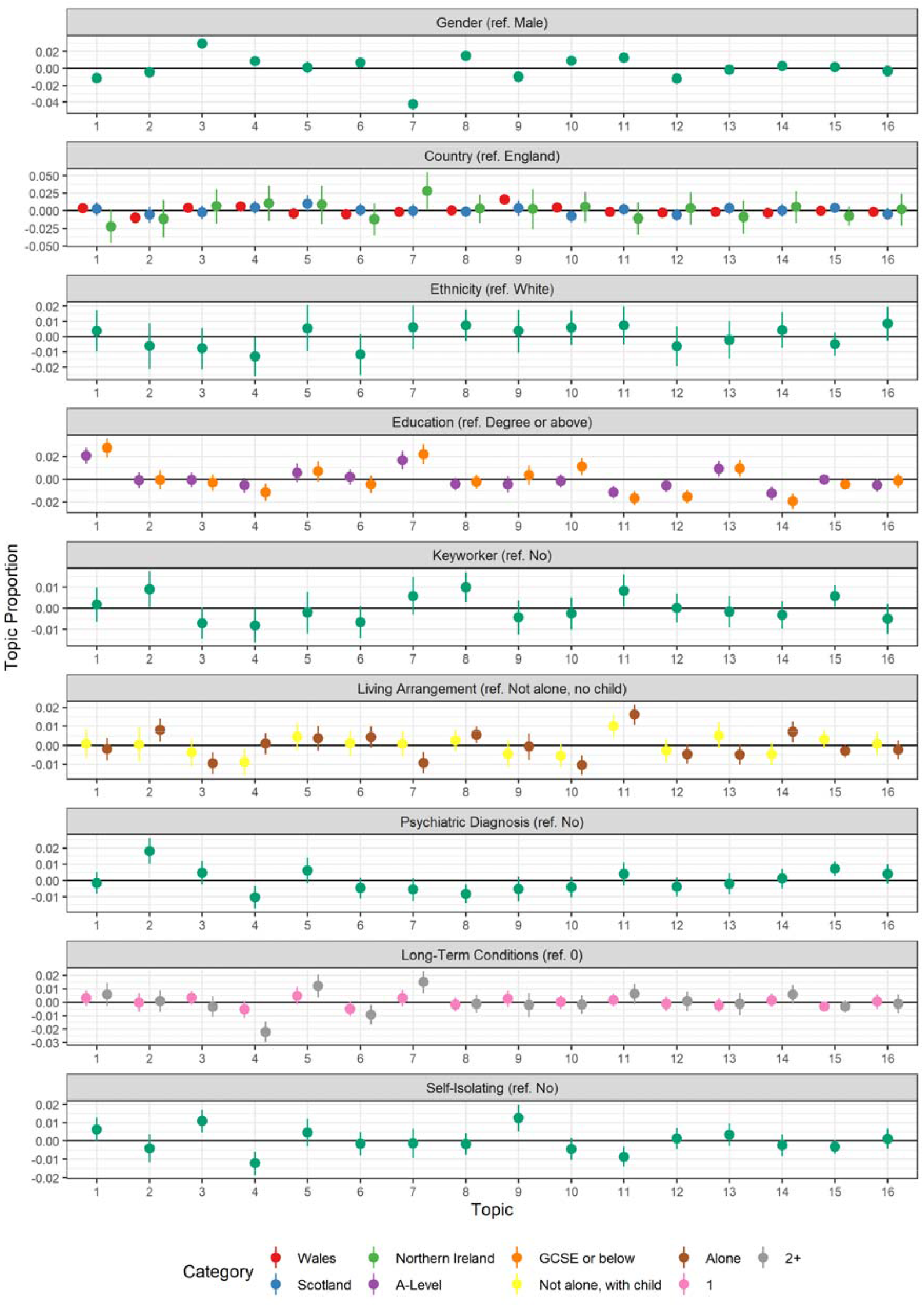
Association between document topic proportion and participants’ demographic and socioeconomic characteristics, health, and confidence in government (+ 95% confidence intervals). Derived from OLS regression models including adjustment for gender, ethnicity, age, education level, living arrangement, psychiatric diagnosis, long-term physical health conditions, self-isolation status, Big-5 personality traits and keyworker status.

### Associations Between Topic Proportions and Lockdown Experiences

The results of regressions assessing the association between lockdown experiences and topic proportions are displayed in Figure 5. Engaging in creative activities (Topic 3), DIY and gardening (Topic 9) and keeping a routine (Topic 12) were associated with greater enjoyment of first lockdown. Creative activities were also related to being excited for future lockdowns or expecting to miss the first lockdown. Following the rules (Topic 7), keeping busy (Topic 10), and thinking positively (Topic 1) were related to being less likely to anticipating missing lockdown. Talking to family and friends was generally related to worse lockdown experiences (though confidence intervals overlapped mean values).

**Figure 5:**
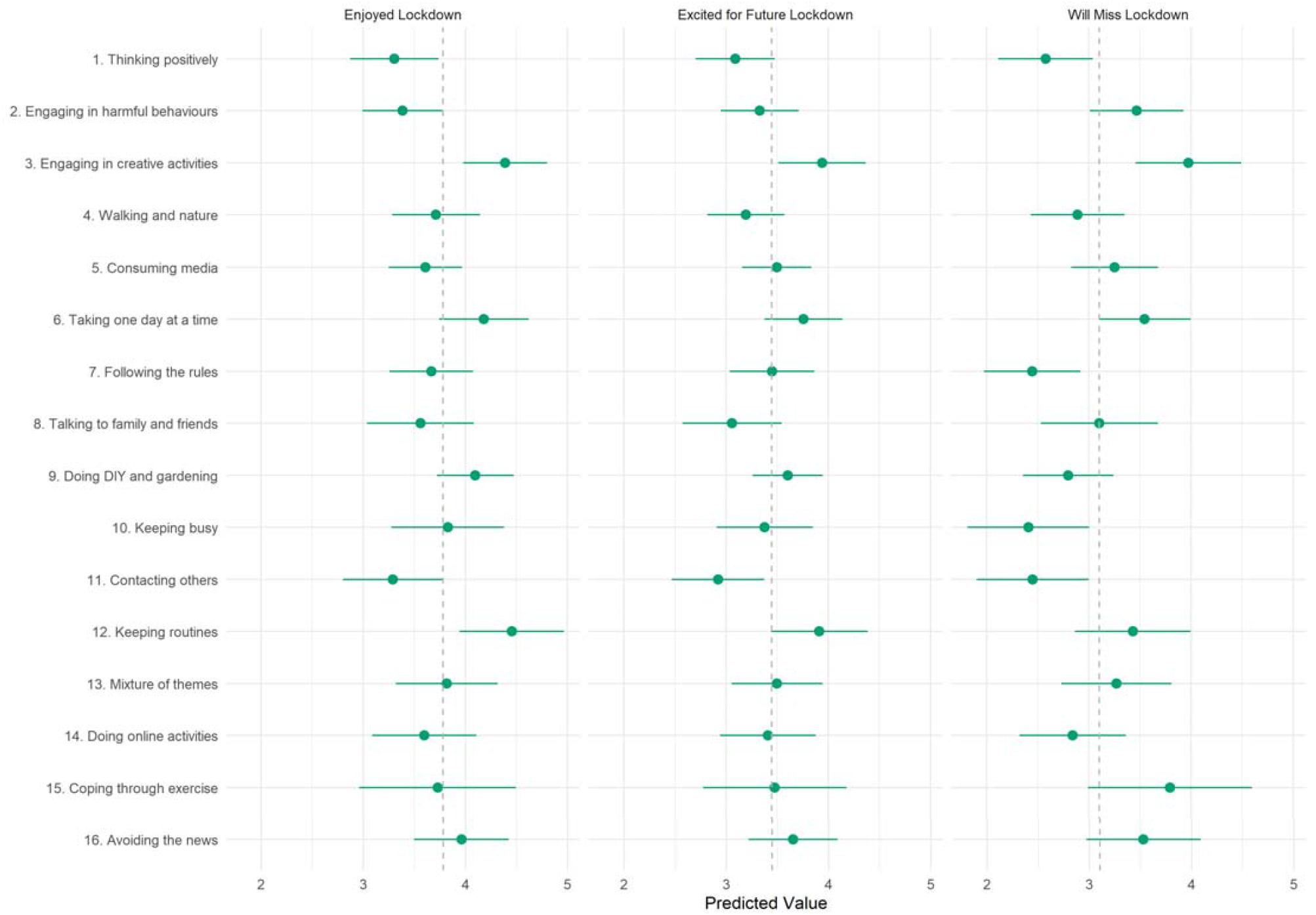
Association between lockdown experiences and document topic proportions (+ 95% confidence intervals).. Dashed line represents mean value for the respective lockdown experience variable

## Discussion

We identified 16 overarching topics of how people were coping during lockdown in the UK. The most discussed coping strategy was ‘thinking positively’ and involved themes of gratefulness and positivity. Numerous topics were centered around activities and hobbies including ‘walking and spending time in nature’, ‘coping through exercise’, ‘doing DIY and gardening’, and ‘engaging in creative activities’. Other themes were digitally oriented, including ‘consuming media’ and ‘doing online activities’, or were socially-supportive, including ‘contacting others’ and ‘talking to friends and family’. Other strategies were more focused on staying in control such as ‘keeping routines’, ‘focusing on one day at a time’, ‘keeping busy’, and ‘following government guidelines’. However, some respondents reported adopting more avoidant strategies including ‘engaging in harmful behaviors’ and ‘avoiding the news’. Coping strategies varied by respondent characteristics including age, personality traits, sociodemographic factors, health characteristics, and lockdown experiences. Individuals who engaged in creative activities or DIY and gardening had the most positive lockdown experiences. Many of the core topics we identified echo those found in other coping research conducted during the COVID-19 pandemic, including reports of ‘embracing lockdown’, feeling hope, and the uptake of numerous hobbies and activities (Ab et al., 2020; Ogueji et al., 2021; Park et al., 2020; Sarah et al., 2021). However, our findings extend this previous research by elucidating specific activities within previous broad themes identified (for example, DIY and gardening). Further, we related these coping strategies to demographic characteristics and experiences of the pandemic.

In line with previous research, a number of known sociodemographic, personality, and health predictors were associated with coping choice during the pandemic (Ahmed et al., 2021; Fluharty & Fancourt, 2021; Park et al., 2020). For example, younger adults in this study reported they were more likely to engage in harmful or risk behaviours; typically older adults are less likely to engage in avoidant coping strategies outside of pandemic circumstances (Chen et al., 2018; Hamarat et al., 2002). There is evidence that older adults experienced generally better mental health across the pandemic than younger age groups (Fancourt, Steptoe, et al., 2021), and in the present study, it is also notable that older adults made more use of keeping busy and doing online activities, all of which may have contributed to their better coping.

Recent evidence suggests socially-supportive strategies (e.g., talking to friends and family, social media, contacting others) have been the most commonly employed coping strategies during the COVID-19 pandemic (Ab et al., 2020; Ogueji et al., 2021; Sarah et al., 2021), and in other infectious disease outbreaks (Chew et al., 2020). Socially-supportive coping during the pandemic has been associated with faster decreased in mental health symptoms during the first lockdown, suggesting it was a particularly effective form of coping (Fluharty et al., 2021). While the most frequently reported strategy in the current study was not socially-supportive (thinking positive), there were two different coping strategies (11.91%) related to a socially-supportive theme (contacting others and talking to friends and family). Previous research has also reported use of such strategies amongst specific populations such as keyworkers and people living alone as ways of maintaining resilience and combatting loneliness, but it was previously unclear whether such usage was above and beyond other demographic groups (May et al., 2021). Our results suggest that not only did these groups use such strategies but they were more likely to do so than non-keyworkers and people not living alone.

The association between creative activities and more enjoyable lockdown experiences is consistent with findings that creative activities are beneficial for a range of mental, social, physical, and wellbeing outcomes (Fancourt et al., 2019). The results are also consistent with a recent analysis of the COVID-19 Social Study suggesting that individual’s used arts activities during lockdown as a way to regulate their emotions (Mak et al., 2021). It is possible that those who were able to engage in creative activities had other advantages, such as higher incomes, larger houses, or fewer caring responsibilities. However, we found no associations with education (a marker of socio-economic status). Further, some of the typical barriers to engagement in the arts changed during lockdown as some activities shifted to a virtual platform (e.g., physical attendance, cost effective) (Mak et al., 2021), which may mean that some people had an enjoyable lockdown experience as they were able to participate in activities they would have otherwise been unable to.

This study had several strengths. We used rich qualitative data from over 11,000 UK adults representing a wide range of demographic groups. By using open-ended free-text data, we were able to analyse spontaneous responses and thus were not limited to coping strategies, activities, or styles we had thought of in advance. Some of the coping strategies were related to participant characteristics in the expected direction – for instance, people with pre-existing mental health conditions were more likely to report engaging in harmful behaviors (in line with previous research that this group is likely to use avoidant coping). This suggests that our models extracted consistent and meaningful themes. While structural topic models are novel in the coping literature, our results show that such models can complement and bridge qualitative and quantitative approaches, providing insights not easily attained with either approach on its own. A further strength of this study was that we used data from 7-8 months after the first lockdown, allowing for an assessment of coping strategies across an extended period of the pandemic.

Nevertheless, this study had several limitations. Not all of the topics identified a single theme consistently and associations with participant characteristics could be driven by idiosyncratic texts. Our sample, though heterogeneous, was not representative of the UK population. Respondents to the free-text question were also biased towards the better educated. This may have generated bias in the topic regression results. While it is plausible that participants discussed coping strategies that they deemed most important, participants may have employed multiple coping strategies and not written about them all. Further, across the long timespan of the pandemic, individuals may have adopted different strategies at different points. Responses may have been biased towards those salient at the time (e.g., those used recently). Moreover, individuals may not interpret or be aware of a behaviour as a coping strategy, though it has that effect – for instance, increasing consumption of alcohol or fatty or sugary foods. A final limitation was that, while we included a wide set of predictors in our models, many relevant factors were unobserved. Associations may be biased by unobserved confounding.

## Conclusions

Sixteen different coping strategies employed by adults in the UK during the COVID-19 pandemic were identified through text-mining participant free test responses to the COVID-19 Social Study. Some strategies reported were more cognitive (or ‘antecedent-focused’), either based around attentional deployment (both focusing attention onto the pandemic by focusing on following the rules or distracting oneself from events by avoiding the news), problem solving (e.g. drawing on social support) or cognitive change (e.g. trying to think more positively about things) (Gross, 2001). Others were response focused, involving the use of hobbies, exercise or substances to cope. Some coping strategies reported were related to more enjoyable lockdown experiences – notably, creative activities. This finding may be useful for helping individuals prepare for future lockdowns or other events resulting in self-isolation. Socially supportive coping also emerged as an important coping strategy used by certain groups at higher risk of poor mental health such as keyworkers and people living alone, highlighting the importance of supporting individuals at risk of increased loneliness and lower social support during pandemics to connect with others. However, more research is needed around potentially maladaptive coping strategies such as harmful and risky behaviours to identify if such behaviours predict poorer mental and physical health during pandemics and how they can be avoided. Overall, this study sheds light onto the important topic of how people adapted to the challenging circumstances of the COVID-19 pandemic and how coping strategies varied by sociodemographic factors. Given that individuals’ roles in pandemics (i.e., survivor, healthcare, patient, caregiver, general population) can also affect how we cope (Chew et al., 2020), future research may want to extend the findings here to explore the interaction between coping strategies, individual roles and subsequent mental health trajectories.

## Data Availability

The free-text data used in this study cannot be made publicly available due to stipulations set out by the ethics committee. The code used in the analysis is available at https://osf.io/xqu8h/.

## Acknowledgements

The researchers are grateful for the support of a number of organisations with their recruitment efforts including: the UKRI Mental Health Networks, Find Out Now, UCL BioResource, HealthWise Wales, SEO Works, FieldworkHub, and Optimal Workshop.

## Author contributions

All authors conceived and designed the study. LW curated the data and conducted the data analysis. LW and MF agreed on narrative titles for the topics and wrote the first draft. All authors provided critical revisions. All authors read and approved the submitted manuscript.

## Data availability

The free-text data used in this study cannot be made publicly available due to stipulations set out by the ethics committee. The code used to run the analysis is available at https://osf.io/xqu8h/.

