## Supplemental Information for "How did people cope during the COVID-19 pandemic? A Structural Topic Modelling Analysis of Free-Text Data from 11,000 UK Adults"

Supplementary Information

### Policy Environment

Management of the COVID-19 pandemic is devolved to the national governments for England (UK Central Government), Scotland, Northern Ireland, and Wales. Nationwide lockdown was first announced in the UK on 23 March 2020, with differences appearing across the home nations in the timing and extent of the release of lockdown restrictions as the UK exited the first wave. The period around 14 October to 26 November overlapped with the beginning of the second wave of COVID-19 in the UK, and there were several key developments in the response to the pandemic over the period, including “circuit-breaker” lockdowns. Figure S1 shows 7-day COVID-19 caseloads and confirmed deaths, along with the Oxford Policy Tracker, a numerical summary of policy stringency (Hale et al., 2020), across the study period. Below, is a summary of the key developments in each country.

#### England

On 12 October, Boris Johnson announced a three-tier system of regional restrictions in England taking effect from 14 October. One area, Liverpool, was assigned the strictest restrictions. On 22 October, increased financial support for jobs and workers was unveiled, with the package announced shortly after areas of the south of England were placed into higher tiers. On 27 October, the UK recorded 367 deaths from COVID-19, the highest daily total since May. Four days later, the government announced a second four-week national lockdown in England, coming into effect from 5 November. Retail and leisure venues closed and individuals were allowed to meet with at most one member of another household in an outdoor setting. The government’s job furlough scheme was extended to the end of March 2021. On 9 November, Pfizer and BioNTech reported effectiveness of 90% in human trials of their COVID-19 vaccine. On 24 November, the four Home Nation governments agree on plan allowing limited household mixing over Christmas. On 26 November, a new tier system was announced for England, with news that many parts of the Midlands and the North of England would be placed in the highest tier when the lockdown ended.

#### Scotland

On 9 October, the Scottish Government imposed restrictions that bars and restaurants in the Central Belt of Scotland (including the high population areas of Glasgow and Edinburgh) must close from 6pm, with the restrictions initially set to last two weeks. On 21 October, the restrictions are extended until 2 November to coincide with the introduction of a new five-tier system of regional restrictions. Most of Scotland is placed in Tier 2, while North and South Lanarkshire, Dundee, and the Central Belt placed in Tier 3. On 20 November, eleven areas in the west of Scotland are placed in tier 4. Bars, restaurants, gyms, and non-essential shops are closed in these areas.

#### Northern Ireland

On 14 October, the Northern Ireland Government announced that, from 16 October, schools would be closed two weeks and to hospitality venues would be restricted takeaway service only for four weeks. On 12 November, restrictions are extended for a further week. On 19 November, the Government announced plans to reimpose restrictions from 27 November, meaning restrictions are eased for a single week.

#### Wales

On 10 October, lockdown restrictions are imposed in Bangor. Six days later, the Welsh Government introduced a travel ban from COVID-19 hotspots across the UK. On 19 October, the Government announced a 17-day circuit-breaker lockdown from 23 October – 9 November. Non-essential shops, pubs, restaurants, and hotels close and supermarkets are advised to sell only essential items during the period. Following the lockdown, two households are allowed to form support bubbles, internal travel restrictions are lifted, and groups of four from different households are allowed to meet indoors. The Government also implements a nationwide system of rules, jettisoning the localised system in place prior to the lockdown.

### Measures

#### Coping free-text question

Coping strategies were measured with a question “What have been your methods for coping during the pandemic so far and which have been the most or least helpful?”, answered via free-text response. This question was asked in data collections between 14 October – 26 November. This question was asked along with seven other free-text questions, which can be see in Table S1.

#### Predictors of topic proportions

To predict topic proportions, we included variables for age, sex, ethnicity, country of residence, education level, living arrangement, keyworker status, self-isolation status, diagnosed psychiatric condition, long-term physical health conditions, and Big-5 personality traits.

Country of residence (England, Scotland, Wales, Northern Ireland), sex (male, female), ethnicity (White, Non-White), age (smooth splines, degrees of freedom 4), education level (GCSE or below, A-levels or equivalent, degree or above), and keyworker status (as working in health, social care or support sectors, or work involving in medicines or PPE production or distribution) were each measured at baseline interview.

Long-term physical health conditions (0, 1, 2+) was measured using a multiple-choice question on medical conditions. Included conditions were high blood pressure, diabetes, heart disease, lung disease, cancer, any other clinically-diagnosed chronic physical health conditions, or any disability. Psychiatric diagnosis (yes, no) was with the same multiple choice question using items on clinically diagnosed depression, clinically diagnosed anxiety, and any other clinically diagnosed mental health problem. Both variables were collected at baseline interview. Self-isolation status was defined as staying at home at any point due to existing medical condition or being categorised as high risk. This variable was collected at data collections between 21 March 2020 – 04 July 2020.

Personality was measured at baseline interview using the Big Five Inventory (BFI-2; Soto & John, 2017), which measures personality on five domains and 15 facets: openness (intellectual curiosity, aesthetic sensitivity, and creative imagination), conscientiousness (organisation, productiveness, and responsibility), extraversion (sociability, assertiveness, and energy level), agreeableness (compassion, respectfulness, and trust) and neuroticism (anxiety, depression, and emotional volatility). Each item was scored on a 5-point scale (1 = “strongly disagree”, 5 = “strongly agree”). We used the sum Likert score for each domain (range 3 - 15). Higher scores indicate higher levels of the trait.

#### Lockdown experiences

Lockdown experiences were collected between 11-18 June 2020 and were measured with three Likert items on enjoying lockdown (How much have you enjoyed lockdown? 1. Not at all, 7. Very much), missing lockdown (Do you feel you will miss being in lockdown? 1. Not at all, 7. Very much), and feelings about future lockdowns (How do you feel about the prospect of any future lockdowns? 1. I would dread it, 7. I would really look forward to it). Note, these questions were asked shortly after the first national lockdown in the UK, 4-5 months prior to the collection of the free-text data and prior to the imposition of new lockdown restrictions in the UK.

### Figures

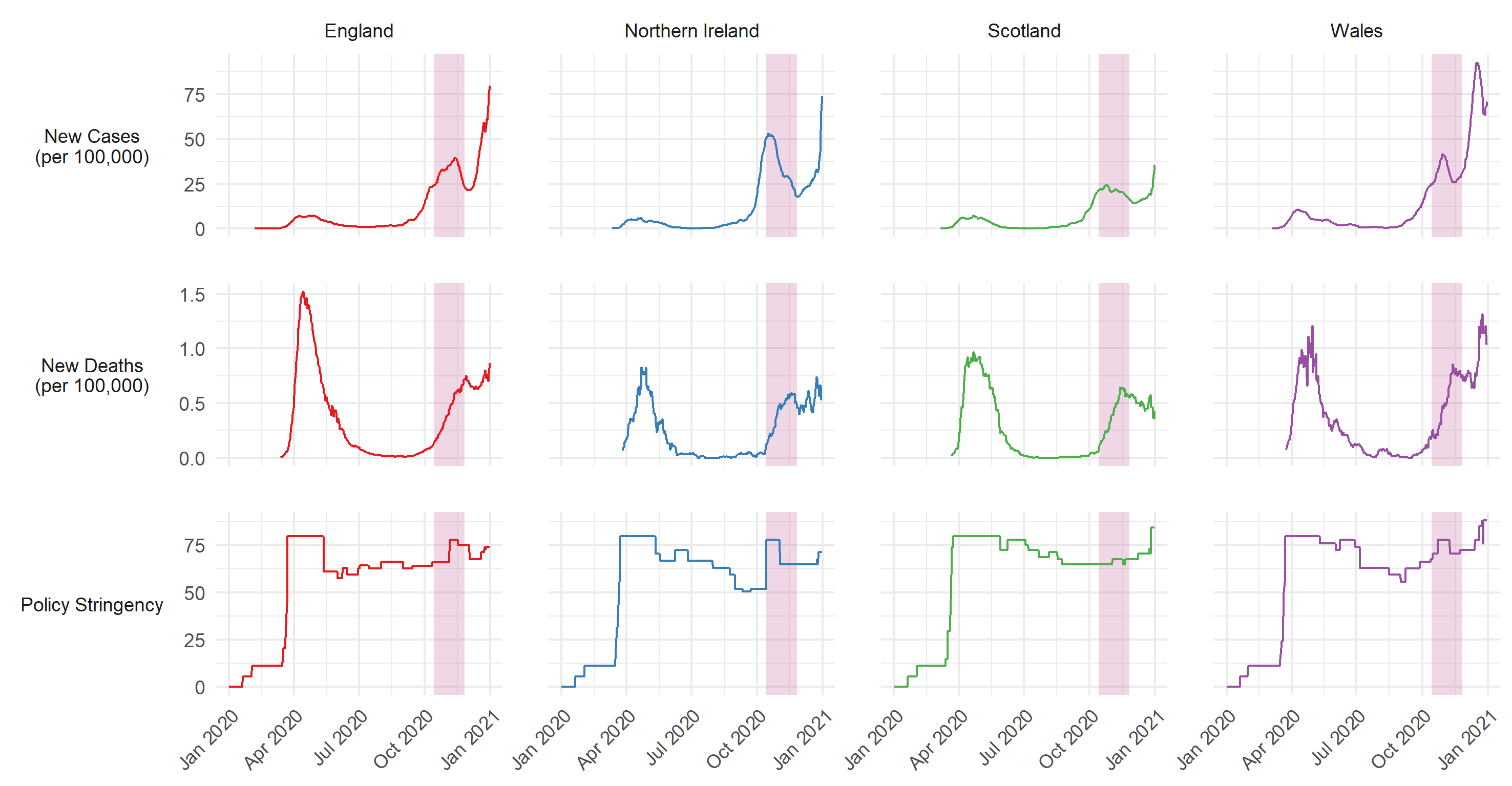

Figure S1: 7-day COVID-19 confirmed cases, COVID-19 deaths, and policy stringency by country. Source: Hale et al. (2020). Pink shaded band represents period in which free-text responses were collected.

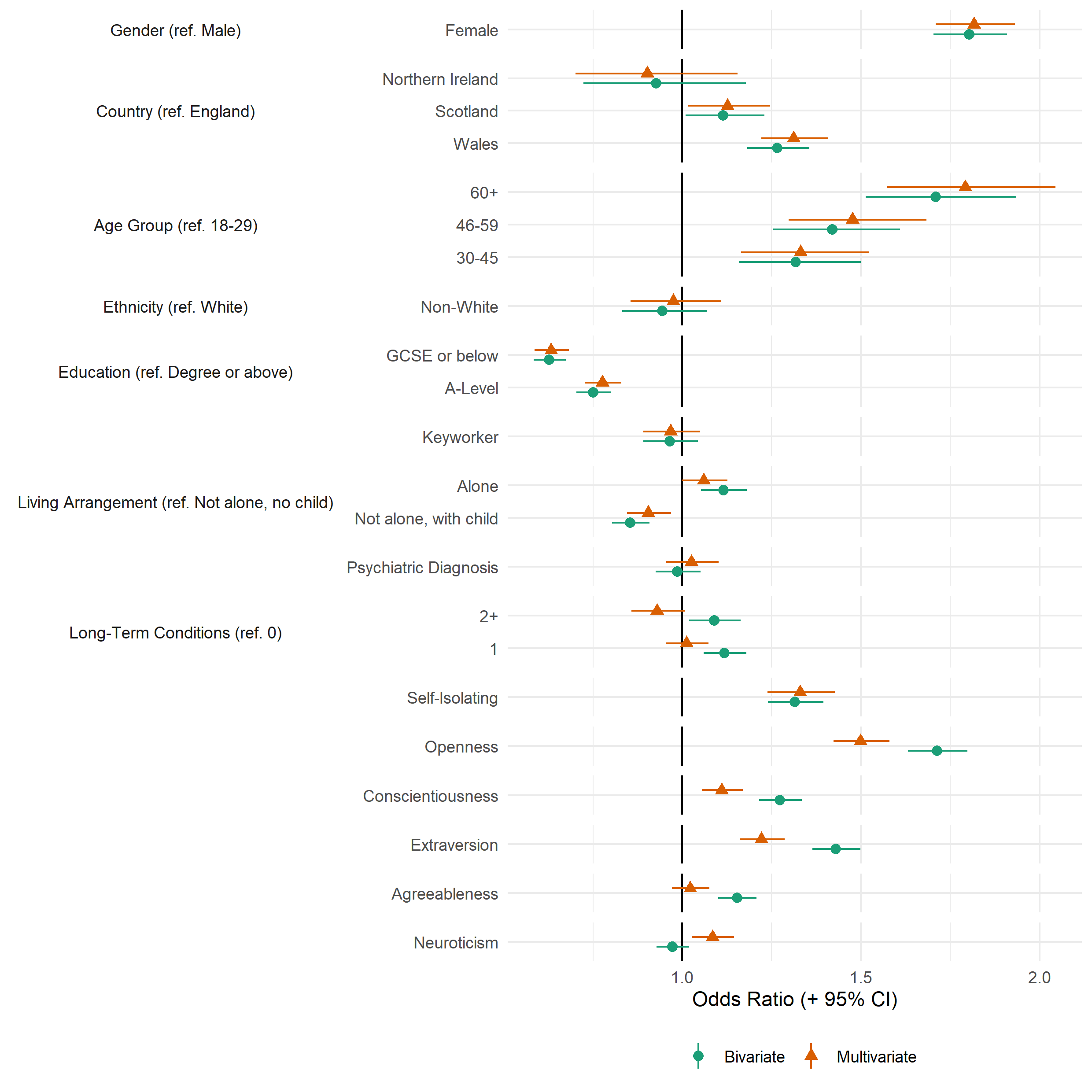

Figure S2: Association between providing a valid free-text response and study variables, derived from bivariate and multivariate logistic regression models.

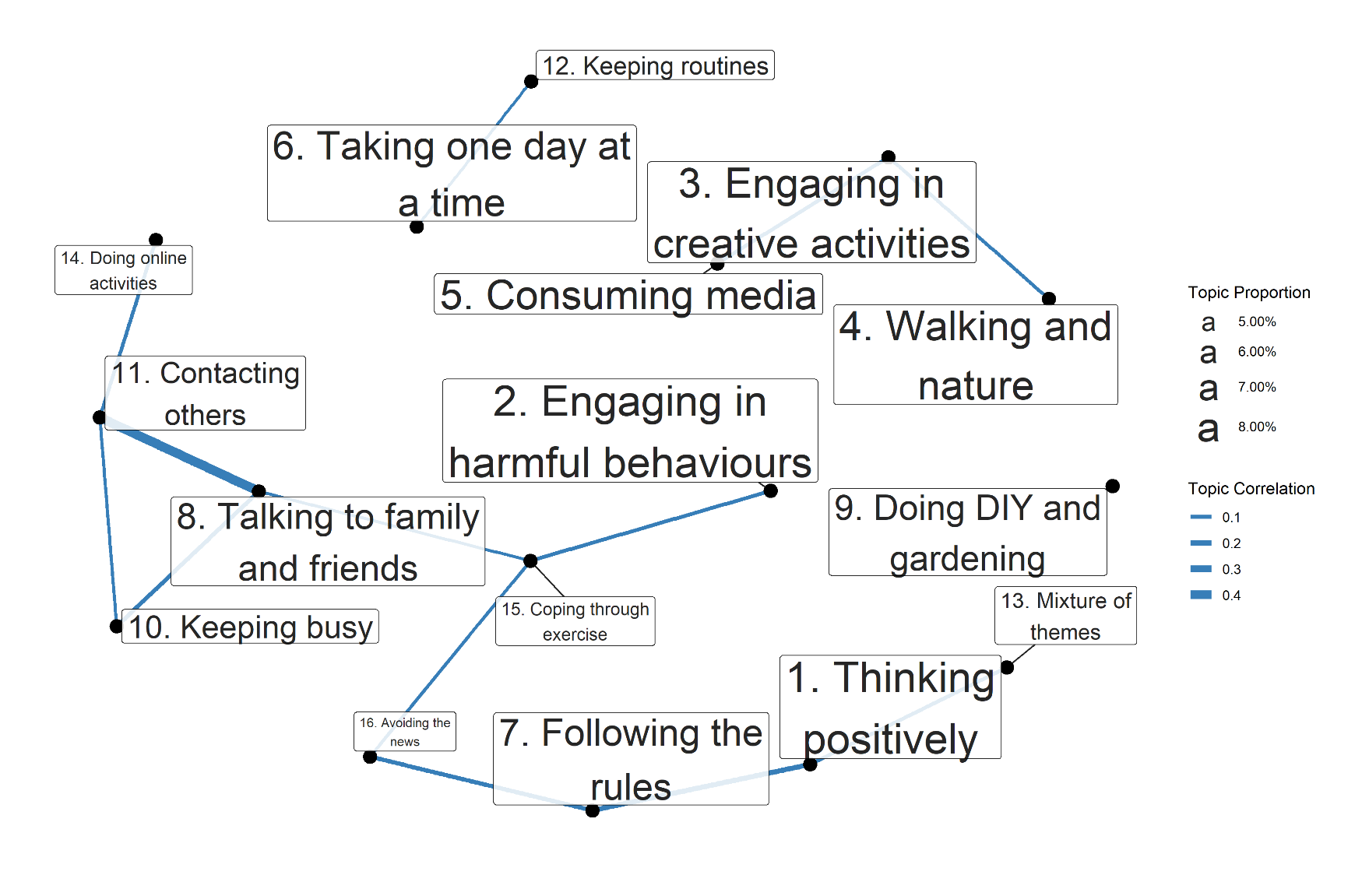

Figure S3: Correlations between estimated topic proportions. Thicker lines indicate greater correlation. A cut-off of ρ = 0.05 is used for the minimum correlations. Labels are sized according to estimated proportion of text devoted to the topic across the free-text responses.

### Tables

Table S1: Free-Text Questions

Question

Q1. Is there anything you would like to tell us about the changes that have been brought about by the Covid-19 pandemic and the impact these have had on your mental health or wellbeing?

Q2. What is bothering you the most about the pandemic? What aspects of it have you been finding most difficult?

Q3. Has the pandemic had any negative impacts on your mental health and wellbeing? If so could you tell us about these?

Q4. Has the pandemic had any positive impacts on your mental health and wellbeing? If so could you tell us about these?

Q5. How have your circumstances (e.g. work, housing, local area, finances, social networks, family life, responsibilities etc ) contributed to your experiences (positive, negative or both) of the pandemic?

Q6. How have your personal attributes (e.g. age, gender, ethnicity, sexuality, health conditions etc) contributed to your experiences (positive, negative or both) of the pandemic?

*Q7. What have been your methods for coping during the pandemic so far and which have been the most or least helpful?

Q8. Since the Covid-19 pandemic began, how have you been feeling about the future? What are you hopeful or concerned about?

* Question used in this analysis

Table S2: Descriptive statistics for outcome variables

|  | Variable | Mean (SD) / n (%) | Missing % |
| --- | --- | --- | --- |
|  | n | 11,073 |  |
| Lockdown Experience | Enjoyed Lockdown | 3.78 (1.72) | 19.9% |
|  | Will Miss Lockdown | 3.1 (1.89) |  |
|  | Excited for Future Lockdown | 3.45 (1.57) |  |
